# The Female Predominant Persistent Immune Dysregulation of the Post COVID Syndrome: A Cohort Study

**DOI:** 10.1101/2021.05.25.21257820

**Authors:** Ravindra Ganesh, Stephanie L Grach, Dennis M. Bierle, Bradley R Salonen, Nerissa M Collins, Avni Y Joshi, Neal Boeder, Christopher V Anstine, Michael R Mueller, Elizabeth C Wight, Ivana T Croghan, Andrew D Badley, Rickey E Carter, Ryan T Hurt

## Abstract

**Objective:** To describe the clinical data from the first 107 patients seen in the Mayo Clinic Post COVID-19 Care Clinic (PCOCC).

**Patients and Methods:** After IRB approval, we reviewed the charts of 107 patients seen between January 19, 2021 and April 29, 2021 in the Mayo Clinic Post COVID Care Clinic (PCOCC) in order to describe the first 107 patients treated through the Mayo Clinic PCOCC. Data was abstracted from the electronic medical record into a standardized database to facilitate analysis. Phenotypes of patients seen in the PCOCC clinic were identified by expert review of predominant symptom clusters.

**Results:** The majority of patients seen in our clinic were female (75%, 80/107), and the median age at presentation was 47 years (interquartile range [IQR] 37, 55). All had Post Acute Sequelae of SARS-CoV-2 infection (PASC) with six clinical phenotypes being identified – fatigue predominant (n=68), dyspnea predominant (n=23), myalgia predominant (n=6), orthostasis predominant (n=6), chest pain predominant (n=3), and headache predominant (n=1). The fatigue-predominant phenotype was more common in women (84%, p=0.006) and the dyspnea-predominant phenotype was more common in men (52%, p=0.002). IL-6 was elevated in 61% of patients (69% of women, p=0.0046) which was statistically discordant with elevation in CRP and ESR which was identified in 17% and 20% of cases respectively (p<0.001). Four PASC phenotypes (fatigue-predominant, myalgia-predominant, orthostasis predominant, and headache-predominant) were associated with central sensitization (CS), and higher IL-6 levels than those phenotypes not associated with CS (p=0.013). Patients with CS phenotypes after COVID-19 infection (post COVID syndrome) were predominantly female (80%, p=0.0085).

**Conclusion:** In our post COVID clinic, we observed several distinct clinical phenotypes. Fatigue-predominance was the most common presentation and was associated with elevated IL-6 levels and female gender. Dyspnea-predominance was more common in men and was not associated with elevated IL-6 levels. IL-6 levels were significantly elevated in patients with PASC and discordant with ESR and CRP, particularly in those with central sensitization phenotypes.

## Introduction

Since its identification in December 2019, the SARS-CoV-2 virus, which causes Coronavirus Disease 2019 (COVID-19), has had a devastating global impact with over 3 million deaths worldwide as of May 2021. During the early COVID-19 pandemic in Italy, a notable proportion of patients who survived the acute illness began presenting with persistent symptoms occurring several weeks after their initial diagnosis. A case series of 143 individuals who had been hospitalized for COVID-19 described persistent symptoms over 60 days from initial onset in 87.4% of cases, showing a predominance of fatigue (53.1%), dyspnea (43.4%), joint pain (27.3%), and chest pain (21.7%) during outpatient follow-up.^1^ A subsequent 6-month cohort study of 1733 patients from Wuhan, China, demonstrated persistent fatigue or muscle weakness (63%), as well as sleep difficulties (26%) and anxiety and depression (23%).^2,^ Recent studies have estimated that between 10-30% of patients who recover from COVID-19 experience persistent symptoms months after resolution of acute illness.^1-7^ These persistent symptoms have been described using several terms including post-COVID syndrome (PoCoS), post-acute COVID-19, and long-haul COVID.^4-7^ Recently the National Institutes of Health (NIH) developed the term Post-Acute Sequelae of SARS-CoV-2 infection (PASC) to describe those COVID-19 patients that have at least one symptom that developed after the acute infection and has persisted after the expected resolution of acute disease.

Direct organ damage from COVID-19 has been observed in PASC patients with single-system involvement such as anosmia, cardiomyopathy, neuropathy, or interstitial lung disease (ILD). Causes of disability beyond lingering effects of direct organ damage include widespread pain, fatigue with post-exertional malaise, orthostatic intolerance, and cognitive impairment including the commonly reported “brain fog”. This is similar to other post infectious syndromes following Epstein-Barr Virus (EBV), Severe Acute Respiratory Syndrome (SARS), Q-fever, Zika, Lyme disease, and Chikungunya. These chronic symptoms resemble those experienced in fibromyalgia (FM), chronic fatigue syndrome (CFS), postural orthostatic tachycardia syndrome (POTS), and central sensitization (CS) syndromes which can often develop in a post-infectious manner. CS syndromes are thought to share common pathophysiology with central neuroinflammation and remodeling of brain and spinal cord pathways leading to enhanced sensitivity to multiple stimuli, sympathetic hyperactivity, and decreased efficacy of inhibitory pathways.^8-19^

Research in CS has recently turned to the role of proinflammatory mediators in the development and persistence of these conditions.^20^ Prominent cytokine and chemokine elevations have included IL-6, IL-8, IL-17, IL-1β, and TNF-α.^20-22^ High sensitivity CRP has been found to be comparably elevated in both fibromyalgia and chronic fatigue syndrome, with mixed data on elevated CRP.^23^ Not surprisingly, there has been similar interest in the cytokine release syndrome attributed to severe cases of COVID-19, especially in regards to IL-6.^24^ The potential overlap between the inflammatory response of CS and PASC has not yet been determined.

In response to the COVID-19 pandemic and the rising cases of PASC, our institution developed a multi-specialty team to coordinate efforts and share clinical and research approaches to PASC. This multispecialty team comprise physicians and scientists from General Internal Medicine (GIM), Preventative Medicine (Prev Med), Physical Medicine & Rehabilitation (PM&R), Psychology, Allergy and Immunology, Infectious Disease (ID), Pulmonology, Neurology, Cardiology, Pediatrics and Otorhinolaryngology (ENT). Subspecialists experienced in acute COVID-19 and PASC see patients with symptoms limited to one organ system, while the GIM (PCOCC) and Prev Med (CARP) clinics evaluate patients with multiple affected systems.^25^

Herein we present our inaugural cohort to describe the clinical presentation and associated laboratory findings from our first 107 patients seen in the Post COVID-19 Care Clinic (PCOCC).

### Patients and Methods

The current study was approved by the Mayo Clinic COVID-19 Research Taskforce and the Mayo Clinic Institutional Review Board. The data for all patients in the PCOCC Clinic and had provided research authorization were entered into a prospectively maintained internal REDCap (Research Electronic Data Capture) database. Patients seen between January 19, 2021 and April 29, 2021 were included. Acute symptoms (within the first 4 weeks of COVID-19 diagnosis or symptom-onset) were recorded, followed by chronic symptoms (those persisting beyond 4 weeks from diagnosis or symptom-onset). Laboratory and other diagnostic testing data (protocol described below) were also collected as available. Data was abstracted and analyzed using R version 3.6.3.^26^

### Care Team Design

Patients who had persistent symptoms post COVID-19 infection were either self- or physician-referred to the PCOCC. All patients completed a standardized questionnaire which contains 52 questions regarding initial COVID-19 infection, symptoms, and treatment, along with ongoing and persistent symptoms that continue to affect them. Patient questionnaires were reviewed by a GIM physician and those with symptoms limited to a single organ system were directly referred to the sub-specialty team for management of their PASC symptoms; for example, patients with anosmia were referred directly to Otorhinolaryngology (ENT). Patients with symptoms in multiple organ systems were initially evaluated via a 30-minute virtual visit for the purpose of introducing the PCOCC clinic and appropriate pre-ordering of tests and consults.

Patients would then be evaluated in person during a condensed itinerary of tests and consults geared toward the predominant symptoms. For many patients, there was no evidence of tissue damage on testing, and these patients likely had a central sensitization (CS) phenotype, including fatigue, myalgia, and orthostasis. For treatment of the CS phenotypes, we created a virtual treatment program aimed at patient education with elements of cognitive behavioral therapy, health coaching, and paced rehabilitation. This program is 8 hours long, delivered as two 4-hour segments, and accompanied by health coaching and nursing follow up for 6 months. As part of the PCOCC evaluation, the team developed a standardized evaluation to be ordered for any patient presenting to the clinic. This included basic laboratory testing (CBC, CMP, ESR, CRP), markers of COVID-19 inflammation (ferritin, D-dimer, IL-6), autoimmune screening (ANA and CRP with reflex to full panel if either positive), 6 minute walk test, and pulmonary function tests and chest imaging if patient had dyspnea.

## Results

### Patient demographics

The first 107 patients evaluated by our PCOCC team were included. Of these, 80 (75%) were female with a median age of 47 years (interquartile range [IQR] 37, 55). 94% of our patients were White/Caucasian, 4% were Black/African American, 2% were American Indian/Pacific Islander, and 2% were unknown. 98% of our patients identified as non-Hispanic and 2% were unknown (Table 1).

**Table 1.** Demographics of patient population treated in PCOCC.

| Characteristic | N = 107<sup>1</sup> |
| --- | --- |
| Age | 47 (37, 55) |
| Sex |  |
| Female | 80 (75%) |
| Male | 27 (25%) |
| Ethnicity |  |
| non-Hispanic | 103 (98%) |
| Hispanic | 2 (1.9%) |
| Unknown | 2 |
| Race |  |
| White | 99 (94%) |
| African/African American | 4 (3.8%) |
| Asian | 0 (0%) |
| Pacific Islander | 0 (0%) |
| American Indian/Native American | 2 (1.9%) |
| Unknown | 2 |

<sup>1</sup>Median (IQR); n (%)

### Phenotypes

Six phenotypes were identified – fatigue predominant (n=68), dyspnea predominant (n=23), myalgia predominant (n=6), orthostasis predominant (n=6), chest pain predominant (n=3), and headache predominant (n=1) (Table 2). Distribution of phenotypes by sex was unequal with more women being predominant for fatigue (84%, *p=0*.*006*), orthostasis (100%), and chest pain (100%). More men were predominant for dyspnea (52%, *p=0*.*002*) and headache (100%). Myalgia-predominant phenotype was evenly distributed among sexes.

**Table 2:** Phenotypes of patients presenting to PCOCC.

| Phenotype | Patients reporting symptom | Women | Men | P-value (Fisher's Exact Test) |
| --- | --- | --- | --- | --- |
| Chest Pain | 3 | 3 (100%) | 0 (0%) | 1 |
| Dyspnea | 23 | 11 (48%) | 12 (52%) | 0.002 |
| Fatigue | 68 | 57 (84%) | 11 (16%) | 0.006 |
| Myalgia | 6 | 3 (50%) | 3 (50%) | 0.17 |
| Orthostasis | 6 | 6 (100%) | 0 (0%) | 0.33 |
| Headache | 1 | 0 (0%) | 1 (100%) | 1 |
| **Total** | **107** | **80** | **27** |  |
| Central Sensitization | 81 | 66 | 15 | 0.008 |

**Table 3:**
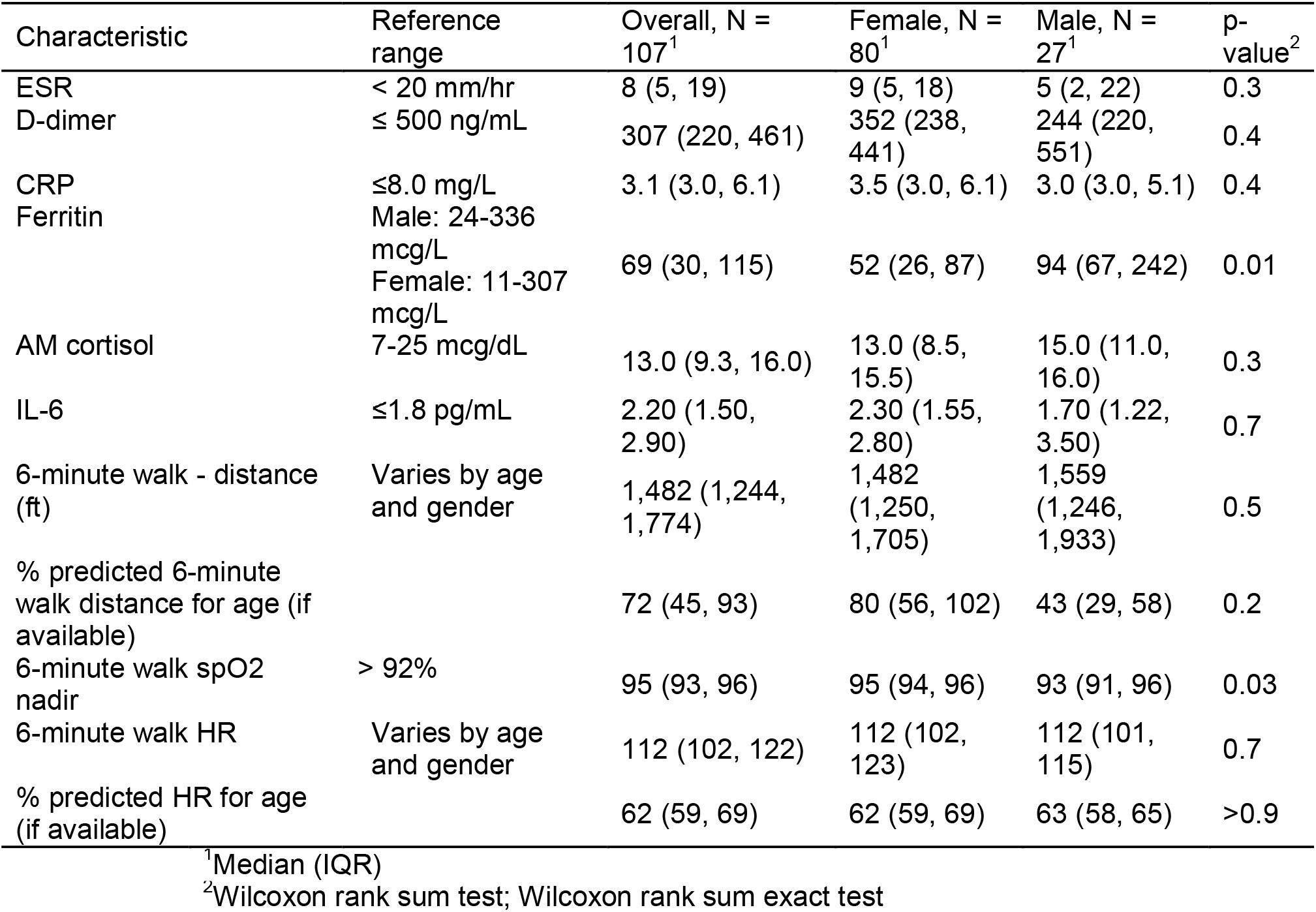
Lab tests by sex.

### Lab markers

Lab markers varied significantly with IL-6 elevated in a higher proportion of patients than CRP (61% vs 17%, p <0.001) and ESR (61% vs 20%, p<0.001) (Figure 1, Table 4). IL-6 was elevated in a statistically significant proportion of women (69% vs 39% in men, p=0.05) while there were no statistically significant differences in elevated ESR (18% women vs 26% in men, p=0.76) and CRP (15% in women vs 21% in men) between the sexes (Table 5). Lab markers also varied by phenotype with IL-6 levels being more elevated in patients with the fatigue-predominant, myalgia-predominant, and orthostasis-predominant phenotypes (Figure 2).

**Table 4.** Inflammatory Markers by Phenotype.

| **Test** | **Fatigue<sup>‡</sup>** | **Dyspnea<sup>‡</sup>** | **Myalgia<sup>‡</sup>** | **Orthostasis<sup>‡</sup>** | **Chest Pain<sup>‡</sup>** | **Headache<sup>†</sup>** |
| --- | --- | --- | --- | --- | --- | --- |
| **IL-6** | 2.5 (1.1, 0.86) | 1.7 (0.9, 0.04) | 4.9 (3.4, 0.0002) | 2.9 (0.3, 0.55) | 1.3 (0.2, 0.27) | - |
| **CRP** | 5.9 (5.2, 0.62) | 4.9 (3.6, 0.47) | 16.04 (25.6, 0.004) | 6.5 (3.6, 0.97) | 3 (0, 0.50) | 34 (0) |
| **ESR** | 13.5 (11.9, 0.46) | 9.1 (7.6, 0.10) | 12.25 (12.2, 0.48) | 21.8 (18.6, 0.05) | 3 (1.4, 0.16) | 42 (0) |

<sup>†</sup>one patient with headache who had an elevated ESR and CRP, IL-6 not measured
<sup>‡</sup>mean (SD, p-value)

**Table 5.** Elevated inflammatory markers by sex.

| **Test** | **Number elevated**<br>**(number with result; %)** | **Women**<br>**(n; %)** | **p-value by Fisher's**<br>**exact test** |
| --- | --- | --- | --- |
| **IL-6** | 41 (67; 61%) | 34 (49; 69%) | 0.046 |
| **CRP** | 12 (71; 17%) | 8 (52; 15%) | 0.72 |
| **ESR** | 14 (69; 20%) | 9 (50; 18%) | 0.76 |

**Table 6.** Characteristics of patients with and without central sensitization (CS)

| **Test** | **CS (n=81)<sup>1</sup>** | **Non-CS (n=26)<sup>1</sup>** | **P-value<sup>2</sup>** |
| --- | --- | --- | --- |
| % female | 81 | 54 | 0.009 |
| Age | 45.6 [13.6] | 48.7 [14.8] | 0.32 |
| IL-6 | 2.75 [1.62] | 1.64 [0.86] | 0.01 |
| CRP | 7.01 [9.67] | 4.55 [3.44] | 0.30 |
| ESR | 14.8 [13.6] | 8.06 [7.49] | 0.05 |

<sup>1</sup>mean [SD]
<sup>2</sup>Fisher exact test (% female); ANOVA (age, IL-6, CRP, ESR)

**Figure 1:**
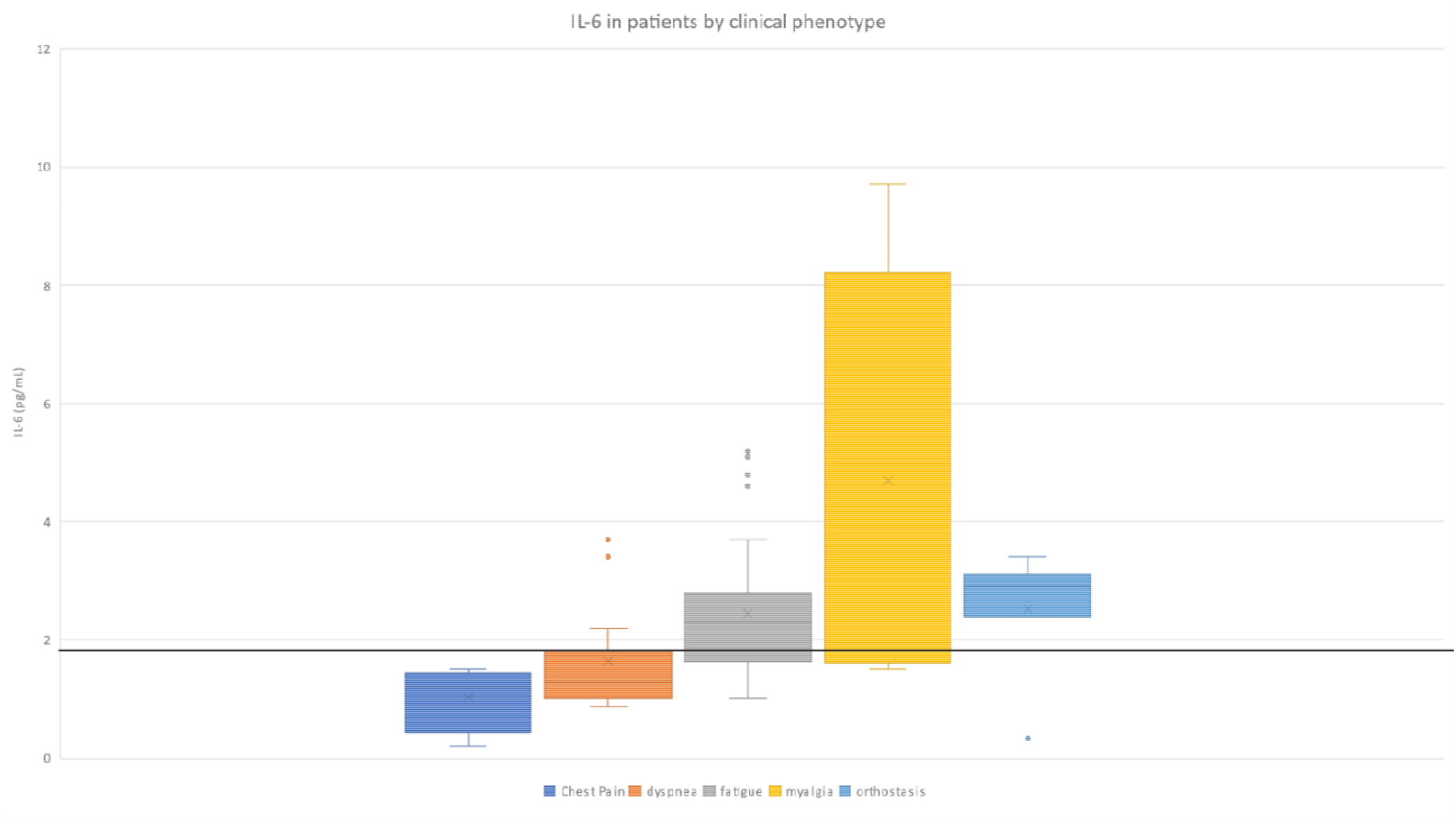
IL-6 in patients by clinical phenotype. Headache predominant phenotype not shown as n = 1 and no IL-6 value for this patient

**Figure 2:**
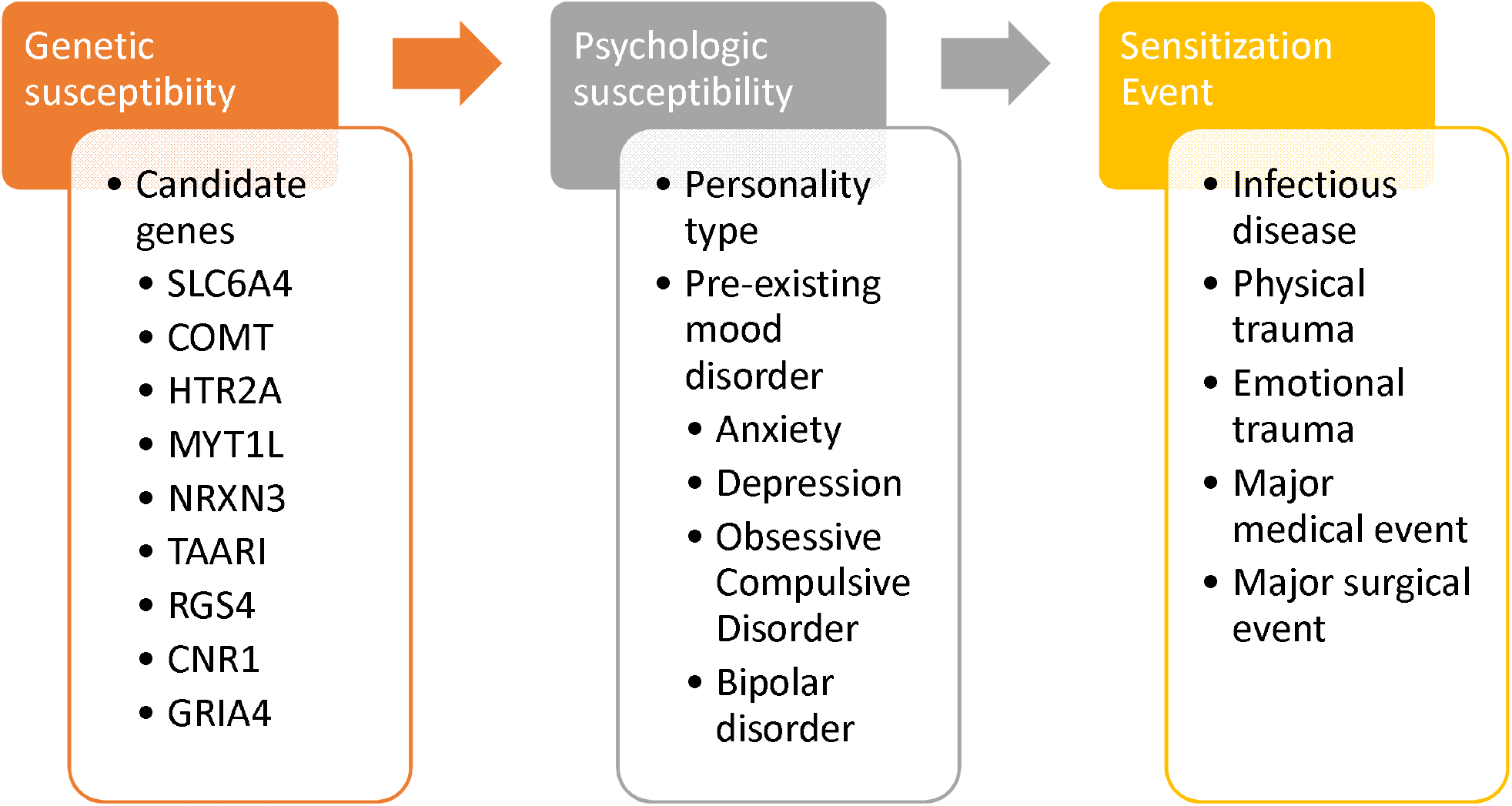
Proposed “three hit” model for development of central sensitization (CS) Candidate genes from D’Agnelli et al, Mol Pain 2019^63^

### Central sensitization

The fatigue-predominant, myalgia-predominant, and orthostasis-predominant phenotypes were considered together as central sensitization phenotypes given their similarity across groups as well as similarity to existing CS conditions of chronic fatigue syndrome, fibromyalgia, and POTS. When considered together against the non-CS phenotype (dyspnea, and chest pain [n=26]), the CS-phenotype (n=81) had statistically significantly higher IL-6 levels (p=0.013) and higher ESR and CRP levels but these did not reach statistical significance (Figure 3, Table 5). The CS-phenotype also had a statistically significantly higher proportion of women (80% vs 27% of men, p=0.0085). Age was not significantly different between patients with CS (mean 45.6, SD 13.6) and without CS (mean 48.7 years, SD 14.8).

## Discussion

The main objective of the PCOCC is to phenotype and then treat PASC patients who are struggling with persistent symptoms for greater than 3 months. There are three main novel findings from the first 107 PASC patients seen in the PCOCC: 1) there is female predominance in the patients seeking care for PASC (n=80; 74.5%) 2) females were more likely to have an elevated IL-6 than males (69% versus 28%) and 3) ongoing fatigue was the most common phenotype in females (n=75;71.3%) and dyspnea most common in males (n=12; 44.4%). Most patients seen in the PCOCC did not have a clear end organ pathology/damage to account for the ongoing symptoms and central sensitization (CS) phenotype (fatigue, myalgia, orthostasis) was the most common categorization. Of patients with central sensitization phenotypes, 66/81 were women (81.4%) consistent with the known female predominance in CS (Table 5).^27, 28^

The majority (n=62; 57.9%) of patients seen in the PCOCC clinic had elevated levels of the pro-inflammatory cytokine IL-6 at least 3 months after acute infection. To our knowledge this has not been described in PASC. Acute COVID-19 infection has demonstrated a similar, but more pronounced pattern of inflammatory response similar to what has been seen in other cytokine release syndromes (CRS) such as sepsis, burns, and acute respiratory distress syndrome. The CRS response includes important components of the innate immune system such as IL-6, IL-8, IL-10 and monocyte chemoattractant (MCP)-1.^29-33^ This innate hyperinflammatory CRS response seen during severe COVID-19 is thought to be the primary mechanism behind adverse outcomes. In addition, IL-6 has been independently associated with mortality in COVID-19 patients requiring mechanical ventilation.^24^ It has been postulated that IL-6 inhibits antiviral activity and blockade has been identified as a potential therapeutic target,^33, 34^ however, recent trials of tocilizumab, a monoclonal antibody against IL-6 receptor alpha in hospitalized COVID-19 patients have shown mixed results.^35, 36^

The present study demonstrated a likely persistent (greater than 3 month) elevated IL-6 level was more frequent in females, which we have hypothesized could be partially responsible for the sex differences being observed in PASC.^37-41^ Sex is one of several variables known to influence the overall immune responses to COVID-19. In a number of acute COVID-19 cohort studies, males had higher rates of hospitalization, critical illness, and mortality when adjusted for comorbidities and age.^38, 41, 42^ A recent cohort study of hospitalized COVID-19 positive patients who had not received immunomodulatory medications and had detailed cytokine, chemokine, and blood cell phenotyping samples compared to healthy controls.^42^ This study found that males had a higher innate inflammatory response (IL-8 and IL-18) than females but when females had elevated innate immune responses it positively correlated with disease progression. Changes in the immune response based on sex do vary throughout the lifecycle with post-puberty/pre-menopausal females having higher levels of inflammatory cytokines (IL-6, TNF, and IL-1β) compared to post-puberty/adult males. These effects may be mediated by sex hormones estradiol, progesterone, and androgens as this difference changes in older age with men having higher inflammatory cytokine levels as they age.^42-44^ The average age of the PCOCC patients was 46.2 years with the average female age being younger than the overall cohort at 44.8 years.

Many patients seen in the PCOCC clinic have clinical presentations very similar to other CS phenotypes such as FMS, CFS and POTS.^45-49^ Symptoms also overlap with the Post ICU Syndrome (PICS) and post-infectious syndromes such as post-treatment Lyme disease syndrome.^45, 50^ This centrally sensitized subset of PASC patients are referred to as having post COVID syndrome (PoCoS). The presence of a common pathway mediated by immune dysregulation provides a common etiology of these conditions and IL-6 is elevated in these syndromes.^51-55^ IL-6 is associated with fatigue and sleep-disturbance in chronic stress and inflammation in central sensitization syndromes.^56^ Treatment of these conditions have traditionally been frustrating as patients will have multiple disabling symptoms yet will have very few (if any) abnormalities on laboratory and procedural testing. The most compelling explanation is that of central sensitization wherein the brain and spinal cord become more sensitive to stimuli thereby reducing stimulus threshold for perception and amplifying existing stimuli.^57^ Changes in the areas of the brain recruited secondary to a standard stimulus vary significantly between healthy controls and those with CS on both PET-CT and functional MRI brain imaging.^58-61^ Besides commonalities in clinical presentation, persistent elevation of immune markers, and changes in functional imaging, patients often display symptoms of persistent sympathetic system hyperactivity with palpitations, exercise intolerance, difficulty initiating and maintaining sleep, and sensitivity to external stimuli.^46-49^ Also seen is a familial preponderance of central sensitization syndromes, with up to a 13-fold increase in incidence of CS compared to the general population.^62^ This suggests a genetic component to CS, which has been supported by several studies identifying genes that occur at a higher frequency in patients with CS, with fibromyalgia being the best studied of these syndromes.^62, 63^ Finally, in our experience, patients affected by these syndromes also share personality traits, often describing themselves as “people pleasers” and “detail-oriented”.^57^ Indeed, there has been some overlay between the genetics of fibromyalgia and anxiety and these conditions are highly comorbid.^64^ With these considerations in mind, we propose a “three hit” hypothesis wherein patients need to have the appropriate candidate genes to have the potential for developing CS, the personality type associated with CS, and then a “sensitizing event” which causes significant systemic distress, often viral in nature, but may include other forms of trauma (life events, physical trauma, surgery, major medical illness) [Figure 4]. The symptoms predominant during the sensitization event often remain as the predominant symptom seen in the illness phenotype but may develop to include other symptoms as the illness progresses. This expansion of symptoms is likely related to secondary changes including impaired sleep and the distress and concern due to the impact of symptoms on life and physical activities.

This study had several limitations. First, the study design is a prospective cohort without strict inclusion criteria which allows for heterogeneity across the sample population. This was intentional as the GIM clinic is best suited for evaluating patients with symptoms across multiple systems. Second, the selection of patients with multiple symptoms increased the concentration of patients with PoCoS, as patients with limited symptoms suggestive of one organ system involvement or those with already identified post COVID end organ damage were directly triaged to the appropriate specialty, e.g. anosmia to ENT, pulmonary fibrosis to Pulmonology.

This was also intentional as we had designed a treatment program directed at helping these individuals. Third, our population was 94% White/Caucasian, and involved no patients of Hispanic descent. This was unintentional and likely secondary to our local demographic and differences in health seeking behaviors. Therefore, future research in a more diverse population is warranted. Fourth, the standard lab panel was not ordered in all patients, which reflects referring provider preference or patient-specific clinical decisions. Because of these limitations, our study data may not be widely generalizable to all PASC patients but provides previously unreported insights into PoCoS and its likely immune dysregulation etiology.

## Conclusions

In our post COVID care clinic, we observed six distinct clinical phenotypes, of which the fatigue and dyspnea-predominant phenotypes were most common in women and men respectively.

The phenotypes resembling known central sensitization syndromes (fatigue-, myalgia-, and orthostasis-predominant) were collectively considered as post COVID syndrome. This phenotype was associated with elevated IL-6, predominantly occurred in women, and presented with three major subtypes – fatigue, myalgia, and orthostasis. Knowledge of these phenotypes and the insights gleaned from the clinical data reported in this report may help in defining etiology and treatment options for PoCoS.

## Data Availability

Data is maintained by authors and available upon request

---

**Supplementary Figure 1:**
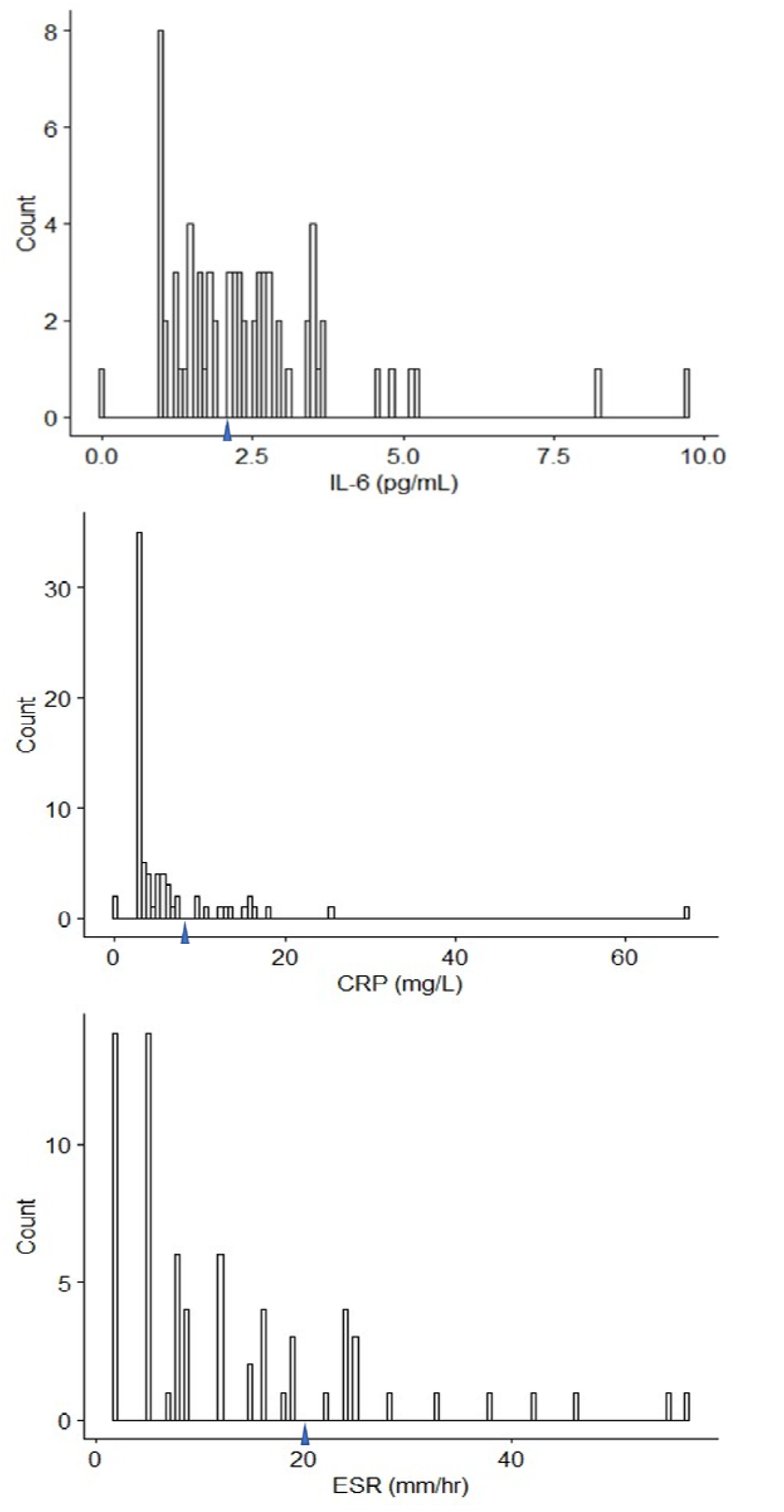
IL-6, CRP, and ESR in patients seen in PCOCC. Blue arrow indicates upper limit of normal for each inflammatory marker

**Supplementary Figure 2:**
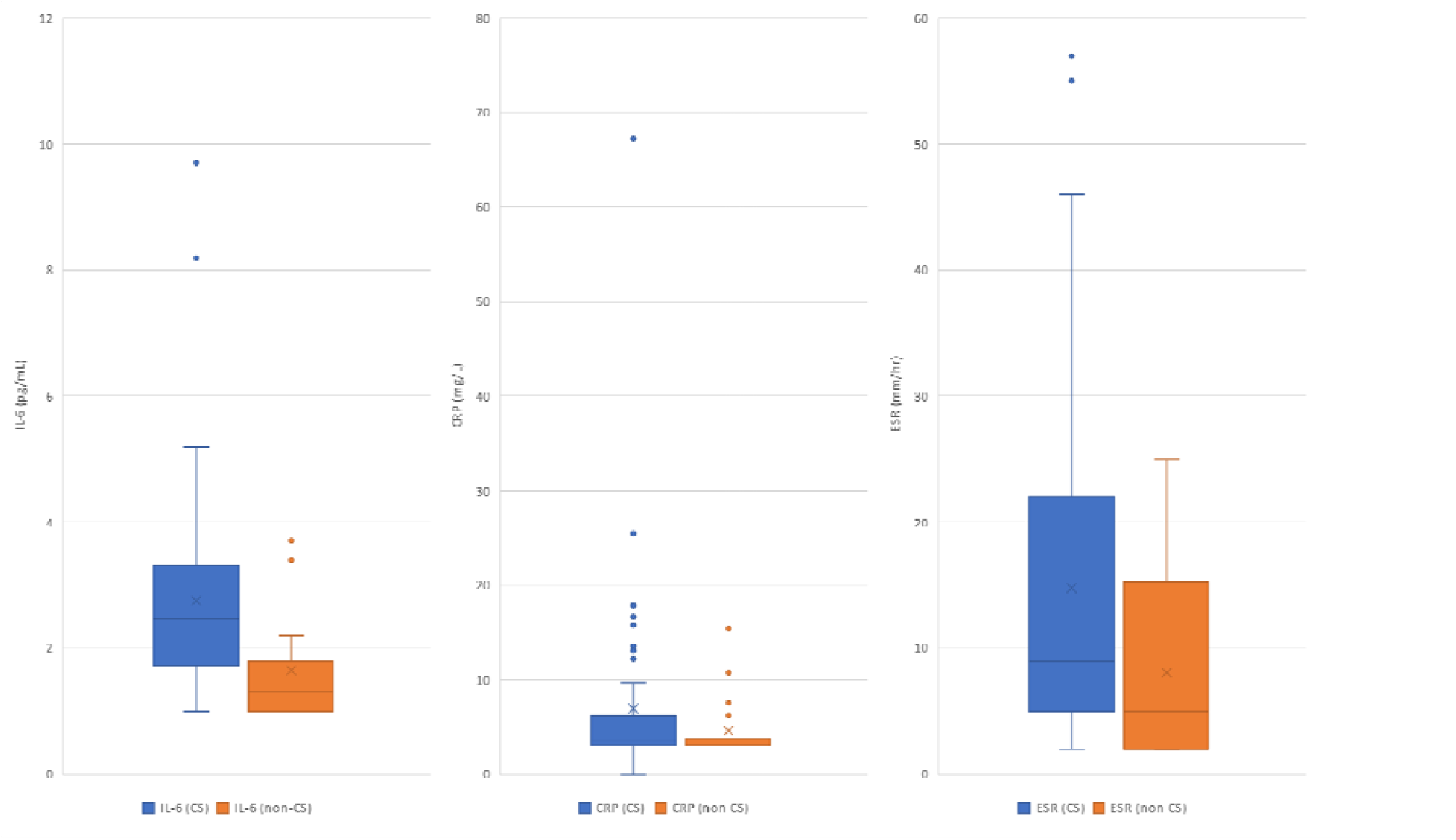
Inflammatory markers in patients with and without central sensitization (CS)

## References

1. CarfÌ A, Bernabei R, Landi F. Persistent Symptoms in Patients After Acute COVID-19. Jama. 2020;324:603–605.

2. Garrigues E, Janvier P, Kherabi Y, et al. Post-discharge persistent symptoms and health-related quality of life after hospitalization for COVID-19. J Infect. 2020;81:e4–e6.

3. Greenhalgh T, Knight M, A’Court C, Buxton M, Husain L. Management of post-acute covid-19 in primary care. Bmj. 2020;370:m3026.

4. Ladds E, Rushforth A, Wieringa S, et al. Persistent symptoms after Covid-19: qualitative study of 114 “long Covid” patients and draft quality principles for services. BMC Health Serv Res. 2020;20:1144.

5. Ladds E, Rushforth A, Wieringa S, et al. Developing services for long COVID: lessons from a study of wounded healers. Clin Med (Lond). 2021;21:59–65.

6. Logue JK, Franko NM, McCulloch DJ, et al. Sequelae in Adults at 6 Months After COVID-19 Infection. JAMA Netw Open. 2021;4:e210830.

7. Townsend L, Dyer AH, Jones K, et al. Persistent fatigue following SARS-CoV-2 infection is common and independent of severity of initial infection. PLoS One. 2020;15:e0240784.

8. Cook RL, Xu X, Yablonsky EJ, et al. Demographic and clinical factors associated with persistent symptoms after West Nile virus infection. Am J Trop Med Hyg. 2010;83:1133–1136.

9. Duvignaud A, Fianu A, Bertolotti A, et al. Rheumatism and chronic fatigue, the two facets of post-chikungunya disease: the TELECHIK cohort study on Reunion island. Epidemiol Infect. 2018;146:633–641.

10. Islam MF, Cotler J, Jason LA. Post-viral fatigue and COVID-19: lessons from past epidemics. Fatigue: Biomedicine, Health & Behavior. 2020;8:61–69.

11. Kristiansen MS, Stabursvik J, O’Leary EC, et al. Clinical symptoms and markers of disease mechanisms in adolescent chronic fatigue following Epstein-Barr virus infection: An exploratory cross-sectional study. Brain Behav Immun. 2019;80:551–563.

12. Leis AA, Grill MF, Goodman BP, et al. Tumor Necrosis Factor-Alpha Signaling May Contribute to Chronic West Nile Virus Post-infectious Proinflammatory State. Front Med (Lausanne). 2020;7:164.

13. Moldofsky H, Patcai J. Chronic widespread musculoskeletal pain, fatigue, depression and disordered sleep in chronic post-SARS syndrome; a case-controlled study. BMC Neurol. 2011;11:37.

14. Pedersen M, Asprusten TT, Godang K, et al. Predictors of chronic fatigue in adolescents six months after acute Epstein-Barr virus infection: A prospective cohort study. Brain Behav Immun. 2019;75:94–100.

15. Pedersen M, Asprusten TT, Godang K, et al. Fatigue in Epstein-Barr virus infected adolescents and healthy controls: A prospective multifactorial association study. J Psychosom Res. 2019;121:46–59.

16. Rodriguez-Morales AJ, Simon F. Chronic chikungunya, still to be fully understood. Int J Infect Dis. 2019;86:133–134.

17. Sejvar JJ, Curns AT, Welburg L, et al. Neurocognitive and functional outcomes in persons recovering from West Nile virus illness. J Neuropsychol. 2008;2:477–499.

18. Sepulveda N, Carneiro J, Lacerda E, Nacul L. Myalgic Encephalomyelitis/Chronic Fatigue Syndrome as a Hyper-Regulated Immune System Driven by an Interplay Between Regulatory T Cells and Chronic Human Herpesvirus Infections. Front Immunol. 2019;10:2684.

19. Shikova E, Reshkova V, Kumanova capital A C, et al. Cytomegalovirus, Epstein-Barr virus, and human herpesvirus-6 infections in patients with myalgic small ie, Cyrillicncephalomyelitis/chronic fatigue syndrome. J Med Virol. 2020.

20. Ji RR, Nackley A, Huh Y, Terrando N, Maixner W. Neuroinflammation and Central Sensitization in Chronic and Widespread Pain. Anesthesiology. 2018;129:343–366.

21. Banfi G, Diani M, Pigatto PD, Reali E. T Cell Subpopulations in the Physiopathology of Fibromyalgia: Evidence and Perspectives. Int J Mol Sci. 2020;21.

22. Theoharides TC, Tsilioni I, Bawazeer M. Mast Cells, Neuroinflammation and Pain in Fibromyalgia Syndrome. Front Cell Neurosci. 2019;13:353.

23. Groven N, Fors EA, Reitan SK. Patients with Fibromyalgia and Chronic Fatigue Syndrome show increased hsCRP compared to healthy controls. Brain Behav Immun. 2019;81:172–177.

24. Popadic V, Klasnja S, Milic N, et al. Predictors of Mortality in Critically Ill COVID-19 Patients Demanding High Oxygen Flow: A Thin Line between Inflammation, Cytokine Storm, and Coagulopathy. Oxid Med Cell Longev. 2021;2021:6648199.

25. Vanichkachorn G, Newcomb R, Cowl CT, et al. Post COVID-19 Syndrome (Long Haul Syndrome): Description of a Multidisciplinary Clinic at the Mayo Clinic and Characteristics of the Initial Patient Cohort. Mayo Clinic Proceedings. 2021.

26. Team RC. R: A language and environment for statistical computing. 2013.

27. Meester I, Rivera-Silva GF, Gonzalez-Salazar F. Immune System Sex Differences May Bridge the Gap Between Sex and Gender in Fibromyalgia. Front Neurosci. 2019;13:1414.

28. Wolfe F, Walitt B, Perrot S, Rasker JJ, Hauser W. Fibromyalgia diagnosis and biased assessment: Sex, prevalence and bias. PLoS One. 2018;13:e0203755.

29. Lorenz G, Moog P, Bachmann Q, et al. Title: Cytokine release syndrome is not usually caused by secondary hemophagocytic lymphohistiocytosis in a cohort of 19 critically ill COVID-19 patients. Sci Rep. 2020;10:18277.

30. Han H, Ma Q, Li C, et al. Profiling serum cytokines in COVID-19 patients reveals IL-6 and IL-10 are disease severity predictors. Emerg Microbes Infect. 2020;9:1123–1130.

31. Ingraham NE, Lotfi-Emran S, Thielen BK, et al. Immunomodulation in COVID-19. The Lancet Respiratory Medicine. 2020;8:544–546.

32. Lagunas-Rangel FA, Chavez-Valencia V. High IL-6/IFN-gamma ratio could be associated with severe disease in COVID-19 patients. J Med Virol. 2020;92:1789–1790.

33. Mazzoni A, Salvati L, Maggi L, et al. Impaired immune cell cytotoxicity in severe COVID- 19 is IL-6 dependent. J Clin Invest. 2020;130:4694–4703.

34. Behm FG, Jin M, Mohapatra G, Barkhordar F, Gavin IM, Gillis BS. Interleukin Deficiency Disorder Patient Responses to COVID-19 Infections. Journal of Advances in Medicine and Medical Research. 2021:40–45.

35. Salama C, Han J, Yau L, et al. Tocilizumab in Patients Hospitalized with Covid-19 Pneumonia. N Engl J Med. 2021;384:20–30.

36. Stone JH, Frigault MJ, Serling-Boyd NJ, et al. Efficacy of Tocilizumab in Patients Hospitalized with Covid-19. N Engl J Med. 2020;383:2333–2344.

37. Huang Y, Pinto MD, Borelli JL, et al. COVID Symptoms, Symptom Clusters, and Predictors for Becoming a Long-Hauler: Looking for Clarity in the Haze of the Pandemic. medRxiv. 2021.

38. Klein SL, Dhakal S, Ursin RL, Deshpande S, Sandberg K, Mauvais-Jarvis F. Biological sex impacts COVID-19 outcomes. PLoS Pathog. 2020;16:e1008570.

39. Klein SL, Flanagan KL. Sex differences in immune responses. Nat Rev Immunol. 2016;16:626–638.

40. Rubin R. As Their Numbers Grow, COVID-19 “Long Haulers” Stump Experts. Jama. 2020.

41. Sudre CH, Murray B, Varsavsky T, et al. Attributes and predictors of long COVID. Nat Med. 2021;27:626–631.

42. Takahashi T, Ellingson MK, Wong P, et al. Sex differences in immune responses that underlie COVID-19 disease outcomes. Nature. 2020;588:315–320.

43. Kragholm K, Andersen MP, Gerds TA, et al. Association between male sex and outcomes of Coronavirus Disease 2019 (Covid-19) - a Danish nationwide, register-based study. Clin Infect Dis. 2020.

44. Raza HA, Sen P, Bhatti OA, Gupta L. Sex hormones, autoimmunity and gender disparity in COVID-19. Rheumatol Int. 2021.

45. Aucott JN, Rebman AW. Long-haul COVID: heed the lessons from other infection-triggered illnesses. The Lancet. 2021;397:967–968.

46. Blitshteyn S, Whitelaw S. Postural orthostatic tachycardia syndrome (POTS) and other autonomic disorders after COVID-19 infection: a case series of 20 patients. Immunol Res. 2021;69:205–211.

47. Poenaru S, Abdallah SJ, Corrales-Medina V, Cowan J. COVID-19 and post-infectious myalgic encephalomyelitis/chronic fatigue syndrome: a narrative review. Ther Adv Infect Dis. 2021;8:20499361211009385.

48. Raj SR, Arnold AC, Barboi A, et al. Long-COVID postural tachycardia syndrome: an American Autonomic Society statement. Clin Auton Res. 2021.

49. Wong TL, Weitzer DJ. Long COVID and Myalgic Encephalomyelitis/Chronic Fatigue Syndrome (ME/CFS)-A Systemic Review and Comparison of Clinical Presentation and Symptomatology. Medicina (Kaunas). 2021;57.

50. Stam HJ, Stucki G, Bickenbach J. Covid-19 and post intensive care syndrome: a call for action. Journal of rehabilitation medicine. 2020;52:1–4.

51. Fletcher MA, Zeng XR, Barnes Z, Levis S, Klimas NG. Plasma cytokines in women with chronic fatigue syndrome. J Transl Med. 2009;7:96.

52. Jason LA, Cotler J, Islam MF, Sunnquist M, Katz BZ. Risks for Developing ME/CFS in College Students Following Infectious Mononucleosis: A Prospective Cohort Study. Clin Infect Dis. 2020.

53. O’Mahony LF, Srivastava A, Mehta P, Ciurtin C. Is fibromyalgia associated with a unique cytokine profile? Rheumatology (Oxford). 2021.

54. Vernino S, Stiles LE. Autoimmunity in postural orthostatic tachycardia syndrome: Current understanding. Auton Neurosci. 2018;215:78–82.

55. Mousa RF, Al-Hakeim HK, Alhaideri A, Maes M. Chronic fatigue syndrome and fibromyalgia-like symptoms are an integral component of the phenome of schizophrenia: neuro-immune and opioid system correlates. Metab Brain Dis. 2021;36:169–183.

56. Rohleder N, Aringer M, Boentert M. Role of interleukin-6 in stress, sleep, and fatigue. Ann N Y Acad Sci. 2012;1261:88–96.

57. Fleming KC, Volcheck MM. Central sensitization syndrome and the initial evaluation of a patient with fibromyalgia: a review. Rambam Maimonides Med J. 2015;6:e0020.

58. Fujii H, Sato W, Kimura Y, et al. Altered Structural Brain Networks Related to Adrenergic/Muscarinic Receptor Autoantibodies in Chronic Fatigue Syndrome. J Neuroimaging. 2020;30:822–827.

59. Maksoud R, du Preez S, Eaton-Fitch N, et al. A systematic review of neurological impairments in myalgic encephalomyelitis/chronic fatigue syndrome using neuroimaging techniques. PLoS One. 2020;15:e0232475.

60. Tirelli U, Chierichetti F, Tavio M, et al. Brain positron emission tomography (PET) in chronic fatigue syndrome: preliminary data. Am J Med. 1998;105:54s–58s.

61. Diers M, Schley MT, Rance M, et al. Differential central pain processing following repetitive intramuscular proton/prostaglandin E(2) injections in female fibromyalgia patients and healthy controls. Eur J Pain. 2011;15:716–723.

62. Arnold LM, Fan J, Russell IJ, et al. The fibromyalgia family study: a genome-wide linkage scan study. Arthritis Rheum. 2013;65:1122–1128.

63. D’Agnelli S, Arendt-Nielsen L, Gerra MC, et al. Fibromyalgia: Genetics and epigenetics insights may provide the basis for the development of diagnostic biomarkers. Mol Pain. 2019;15:1744806918819944.

64. Fernandez-de-Las-Penas C, Ambite-Quesada S, Gil-Crujera A, Cigaran-Mendez M, Penacoba-Puente C. Catechol-O-methyltransferase Val158Met polymorphism influences anxiety, depression, and disability, but not pressure pain sensitivity, in women with fibromyalgia syndrome. J Pain. 2012;13:1068–1074.

